# Ruminant-dense environments increase risk of Shiga toxin-producing *Escherichia coli* independently of ruminant contact

**DOI:** 10.1101/2024.09.25.24314361

**Authors:** Caitlin Ward, William Finical, Kirk Smith, Joshua M. Rounds, Carrie A. Klumb, Gillian A.M. Tarr

## Abstract

Cattle and other domestic ruminants are the primary reservoirs of O157 and non-O157 Shiga toxin-producing *Escherichia coli* (STEC). Living in areas with high ruminant density has been associated with excess risk of infection, which could be due to both direct ruminant contact and residual environmental risk, but the role of each is unclear. We investigated whether there is any meaningful risk to individuals living in ruminant-dense areas if they do not have direct contact with ruminants. Using a Bayesian spatial framework, we investigated the association between the density of ruminants on feedlots and STEC incidence in Minnesota from 2010 to 2019, stratified by serogroup and season, and adjusting for direct ruminant contact. For every additional head of cattle or sheep per 10 acres, the incidence of O157 STEC infection increased by 30% (IRR 1.30; 95% CrI 1.18, 1.42) or 135% (IRR 2.35; 95% CrI 1.14, 4.20), respectively, during the summer months. Sheep density was also associated with O157 STEC risk during winter (IRR 4.28; 95% CrI 1.40, 8.92). The risk of non-O157 STEC infection was only elevated in areas with goat operations during summer (IRR 19.6; 95% CrI 1.69, 78.8). STEC risk associated with ruminant density was independent of direct ruminant contact across serogroups and seasons. Our findings demonstrate that living in a ruminant-dense area increases an individual’s risk of O157 and non-O157 STEC infection even without direct ruminant contact, indicating that prevention efforts need to extend to community strategies for averting indirect transmission from local ruminant populations.

**Importance:** STEC are zoonotic enteric bacteria responsible for 2.5 million illnesses each year. Infections in young children can be especially devastating, causing hemolytic uremic syndrome (HUS), a debilitating and sometimes fatal form of acute kidney injury. STEC’s primary reservoirs are cattle and other domestic ruminants, and transmission can occur through food, water, animal contact, and person-to-person. Living near ruminants poses a significant risk of STEC infection; however, the proportion of that risk due to direct ruminant contact or other routes of transmission is unknown. Our research demonstrates that direct ruminant contact is a substantial risk irrespective of location, and that individuals living in ruminant-rich regions are at high risk of STEC infection regardless of whether they come into contact with ruminants. These findings indicate a need for multi-pronged prevention efforts that emphasize control of contamination in the environments surrounding ruminant populations, in addition to biosafety precautions when contacting ruminants directly.

## Introduction

Shiga toxin-producing *Escherichia coli* (STEC) are estimated to cause 2.5 million illnesses each year (1), including 265,000 in the United States (2). *E. coli* O157:H7 remains the single most common serotype in the U.S., but reported infections with non-O157 STEC surpassed infections with the O157 serogroup in 2013 (3).

Cattle are considered STEC’s primary reservoir, but STEC have been isolated from a wide variety of species, and small ruminants such as sheep and goats have also been recognized as important reservoirs (4). STEC can be transmitted from their animal reservoirs to human populations through food, direct animal contact, and contaminated environments, including water. While the largest STEC outbreaks are predominantly due to nationally or internationally distributed food products, the majority of STEC cases are sporadic (5).

The role of local reservoirs in an individual’s risk of STEC infection is unclear. Reported cases are disproportionately from rural populations and individuals with animal contact. In Scotland, an estimated 26% of O157 STEC infections were of environmental origin due to livestock in rural areas (6). In Minnesota, 22% of O157 and 16% of non-O157 reported STEC infections had an animal agriculture exposure, and large proportions specifically had contact with ruminants or ruminant environments (7). Concordantly, multiple studies have identified local cattle density as a risk factor for STEC infection (8–13), and the density of small ruminants has also been associated with increased risk of STEC infection (6, 10, 14). One study examining the effect of cattle, sheep, and goat density on STEC risk found that only goat density was associated with incidence (14), suggesting that some of the studies identifying cattle density as a risk factor could be confounded by small ruminant density. Additionally, the majority of studies on ruminant density have focused exclusively on the O157 serogroup. One study that examined multiple STEC serogroups found differences in the presence and magnitude of the effect of cattle density on incidence (11), supporting arguments that reservoirs and transmission for O157 and non-O157 STEC should not be assumed to be the same (15).

The existing studies on livestock density have been conducted almost exclusively in Europe. Geographic variation in STEC strains has been well-established (16, 17), and there is evidence for genetic separation of strains from different hosts, driven by geography (18), suggesting the potential for host differences between Europe and North America. Most importantly, current studies of ruminant density have not separated the effects of direct animal contact and residual environmental risk. It is unknown whether the risk associated with ruminant density is primarily due to direct animal contact, a known risk factor (19–22), or whether simply living in an area with ruminants significantly increases an individual’s risk of STEC infection even in the absence of direct contact.

Here, we undertake one of the first investigations of the role of ruminant density on O157 and non-O157 STEC incidence in North America. We include cattle, sheep, and goat density in our analysis to examine the effect of and simultaneously adjust for the collocation of multiple species. In the first study to combine ruminant density and direct animal contact, we use direct contact exposure information reported by cases to adjust for direct ruminant contact, isolating the effect of ruminant density. We find that cattle and sheep density are associated with O157 incidence, goats are strongly associated with non-O157 incidence, and the risk posed by ruminant density is independent of direct ruminant contact. Our findings demonstrate that living in a ruminant-dense area is sufficient to increase an individual’s risk of O157 and non-O157 STEC infection even without direct ruminant contact, indicating that prevention efforts need to extend to community strategies for averting indirect transmission from local ruminant populations.

## Materials & Methods

### Study Population

Laboratory-confirmed (23), symptomatic STEC cases reported to the Minnesota Department of Health (MDH) from 2010 through 2019 with complete data for home zip code were included in the analysis. MDH conducts statewide, active surveillance for STEC. Individuals with STEC were asked if they lived on, worked on, or visited a farm and if they had visited a public animal venue such as a petting zoo, fair, or educational exhibit. Those responding yes were asked about the presence of and direct contact with different animals. Reported contact with cattle, sheep, or goats during the 7 days prior to illness were combined into a single direct ruminant contact variable. We defined presence of cattle, sheep, or goats without direct contact, which constituted exposure to the ruminant environment, as indirect ruminant contact. To stratify by season, November through April were defined as “winter” and May through October as “summer”. Missing data for age, sex, direct and indirect ruminant contact, and serogroup were imputed using the MICE package in R (24, 25). We imputed 40 datasets with 5 iterations each. Because we used a Bayesian approach for the primary analysis, we summarized the imputed data in a single dataset using the mode of each imputed variable for each individual. This study was ruled exempt by the University of Minnesota IRB.

The 2010 census was used to obtain age/sex stratified population counts at the zip code tabulation area (ZCTA) level (26). Age categories were collapsed to 0-4, 5-9, 10-49, and ≥50 years to reflect important age populations for STEC infection. Sex was classified as male or female, resulting in eight total age/sex categories.

Minnesota ZCTA boundaries were obtained from the Minnesota Geospatial Commons (27). The data contained boundaries for 887 ZCTAs fully or partially in the state of Minnesota. Of these, six ZCTAs specific to a single business or business block were not contained in the 2010 Census data and were dropped, yielding 881 ZCTAs for final analysis.

### Food Production Animals

Data on food animal production facilities in Minnesota are reported to the Minnesota Pollution Control Agency and were obtained from the Minnesota Geospatial Commons (January 27, 2023) (28). Per state law, the Agency defines and registers all operations capable of holding ≥50 animal units, or ≥10 animal units within shorelands as “feedlots”. Data included latitude/longitude, registration information, and animal counts and types for 25,062 feedlot facilities with animals. Data were updated daily and contained only the most current registration information for each feedlot, with 4.7% of feedlots having new registrations or permit issuances, 94.7% having updated registrations, and 0.5% with missing registration types. Of all new registrations, 99% had registration periods of 4 years or more. Based on this information, feedlots with new registration dates during the period 2010-2019 or updated registrations within 4 years of 2019 (2020–2023) were retained for analysis (n=24,410). Animal types considered for analysis were cattle (including dairy and beef cattle, bulls, and veal calves), swine, goats, and sheep. Latitude and longitude coordinates of each feedlot were used to identify their ZCTA, and the total number of animals in each ZCTA was calculated. Animal densities were calculated by dividing the total number of animals by the area in acres of each ZCTA and multiplying by 10 to yield the number of animals per 10 acres.

### FoodNet Population Survey

While contact with ruminants was ascertained directly from STEC cases, ruminant contact in the rest of the population was unknown but was needed to establish an expected number of individuals with ruminant contact. We determined the number of individuals per ZCTA expected to have ruminant contact using the most recent CDC FoodNet Population Survey, which was administered in 2018-2019 (29). The survey ascertained information on acute diarrheal illness and associated risk factors, including whether an individual had contact with cattle, sheep, or goats during the past 7 days. Demographics on each survey respondent were also collected, including ZCTA and county of residence, age, sex, and month of response. As not all ZCTAs were represented in the survey, we conducted subsequent analyses at the county level. We mapped ZCTAs to counties using the Census ZCTA to County Relationship File (30) by assigning the largest Minnesota county by area that overlapped the zip code. Age, sex, and month of response were completed for all respondents. Individuals with missing county of residence or animal contact were excluded from the analysis.

Three sampling frames (landline phone, cell phone, and address-based (ABS)) and two data collection modes (computer-assisted telephone (CATI) and web) were used for data collection. To account for systematic differences in responses and estimates across the sampling frames and data collection modes, a collapsed variable was created with three categories: landline/CATI, cellphone/CATI, and ABS/Web.

### Modeling the Expected Population with Animal Contact

We used the FoodNet Population Survey data in a Bayesian spatial model to estimate the probability of contact with cattle, goats, or sheep across the state. For the 87 counties in the state of Minnesota, boundaries were obtained from the Minnesota Geospatial Commons (February 22, 2023) (31). The recommended analysis approach for the FoodNet Population Survey data uses weighted generalized estimating equations (GEE) to fit a marginal model while controlling for correlation across the three mode/frame categories. While GEE marginal estimation does not translate to the Bayesian disease mapping framework, we incorporated mode/frame in the analysis. The number of respondents in each county that reported contact with ruminants was assumed to be binomially distributed with size equal to the total number of respondents in the county. The probability of contact was modeled using a logit link function and covariates for age group, season, and mode/frame category; sex was initially also included but removed to reduce variability in the analysis. The estimates were collapsed across mode/frames by a weighted average approach. The model also included a spatial random effect using an intrinsic Conditional Autoregressive prior and an uncorrelated heterogeneity term to account for potential overdispersion, which was modeled using an independent and identically distributed normal prior. Independent, diffuse normal priors were used for coefficients of the fixed effects. The model was fit using the R package NIMBLE (32, 33). Convergence was assessed on three independent chains with different starting values using the Gelman-Rubin diagnostic (34), with all parameters having values below 1.1.

Using the estimated county-level posterior mean probabilities, we calculated the expected number of ruminant contacts in each ZCTA in each season by age and sex. To convert from the county level to ZCTA level, all ZCTAs within a county were assumed to have the same expected probability. Then, the expected probability of ruminant contacts in summer and winter were multiplied by the ZCTA population in each age/sex group yielding an expected number of contacts for both seasons by strata. Subtracting the expected number of contacts from the total population, we also calculated the expected number of individuals with no ruminant contact. In summary, we split the total population in each ZCTA into 16 strata based on the four age groups, two sexes (although the estimated probability of contact was constant by sex), and expected ruminant contact in the past 7 days.

### Modeling STEC Incidence by Ruminant Density and Contact

The outcome of interest was the number of STEC cases in each ZCTA during the period 2010-2019. As this is a count outcome, a Poisson model with a log link was used. Separate models were fitted by season (summer: May-October, winter: November-April) and by STEC serogroup (O157 vs. non-O157). Across all models, an offset for the total person-years at risk in each age/sex/contact strata and ZCTA was used. Two groups of models were created, one without individual-level direct ruminant contact and one incorporating a binary covariate for direct ruminant contact for each case. In both models, cattle, sheep, and goat density were included as covariates.

As before, a Bayesian disease mapping approach was used with spatially structured and uncorrelated random effects. Independent, diffuse normal priors were used for coefficients of the fixed effects. Computation was done using NIMBLE (32, 33) with convergence of three chains assessed using the Gelman-Rubin diagnostic (34), with all parameters having values below 1.1. From the models with direct contact included, we estimated the age-sex-contact-adjusted STEC rates per 100,000 individuals in each ZCTA. These adjusted rates were computed by combining the age-sex-contact-specific estimated STEC rates across age/sex/contact groups via direct standardization. Covariate effects were expressed as incidence rate ratios (IRRs) with 95% Bayesian credible intervals (CrI). Incidence rates were mapped using cutoffs derived from the Healthy People 2020 goal for O157 STEC incidence of 0.6/100,000 and Healthy People 2030 goal for all STEC incidence of 3.7/100,000 (35, 36).

### Sensitivity Analysis

To assess the potential role of swine in STEC incidence in Minnesota, we repeated the primary analysis including swine density. We also assessed whether incorporating indirect exposure to ruminants (i.e., exposure to ruminant environments), in addition to direct contact, meaningfully affected our results.

## Results

There were 3,048 symptomatic STEC cases reported to MDH from 2010-2019. Of these, a valid zip code was unavailable for two, resulting in 3,046 cases for analysis. STEC serogroup was identified for 2,905 (95%) infections, including 1,225 (40%) O157 and 1,680 (55%) non-O157 STEC (Table 1). Overall, cases were more likely to be female (59%), and 608 (20%) were <5 years old. A slightly greater proportion of O157 than non-O157 STEC cases were reported during the summer months (78% vs. 66%) and reported direct contact with ruminants (10% vs. 7.5%). Incidence of STEC differed substantially by ZCTA, with most areas falling at the extremes with <0.6 cases per 100,000 or >7.4/100,000 (Figure 1).

**Table 1.** Characteristics of reported STEC cases, Minnesota 2010-2019, by serogroup. Unknown values were imputed prior to analysis.

| **Characteristic** | **O157**<br>**N=1,225** | **Non-O157**<br>**N=1,680** | **Unclassified**<br>**N=141** | **Overall**<br>**N=3,046** |
| --- | --- | --- | --- | --- |
| **Age (years)** | 26.77 (24.25) | 27.62 (22.56) | 30.52 (24.20) | 27.41 (23.34) |
| **Unknown** | 0 | 0 | 1 (0.7%) | 1 (<0.1%) |
| **Age group** |  |  |  |  |
| **0-4** | 268 (22%) | 313 (19%) | 26 (19%) | 607 (20%) |
| **5-9** | 126 (10%) | 96 (5.7%) | 8 (5.7%) | 230 (7.6%) |
| **10-49** | 565 (46%) | 946 (56%) | 75 (54%) | 1,586 (52%) |
| **50+** | 266 (22%) | 325 (19%) | 31 (22%) | 622 (20%) |
| **Unknown** | 0 | 0 | 1 (0.7%) | 1 (<0.1%) |
| **Gender** |  |  |  |  |
| **Female** | 670 (55%) | 1,030 (61%) | 84 (60%) | 1,784 (59%) |
| **Male** | 554 (45%) | 648 (39%) | 56 (40%) | 1,258 (41%) |
| **Unknown** | 1 (0.1%) | 2 (0.1%) | 1 (0.7%) | 4 (0.1%) |
| **Contact with ruminants** |  |  |  |  |
| **Yes** | 124 (10%) | 126 (7.5%) | 6 (4.3%) | 256 (8.4%) |
| **No** | 705 (58%) | 925 (55%) | 6 (23%) | 1,662 (55%) |
| **Unknown** | 396 (32%) | 629 (37%) | 103 (73%) | 1,128 (37%) |
| **Season** |  |  |  |  |
| **Summer** | 955 (78%) | 1,117 (66%) | 103 (73%) | 2,175 (71%) |
| **Winter** | 270 (22%) | 563 (34%) | 38 (27%) | 871 (29%) |

**Figure 1.**
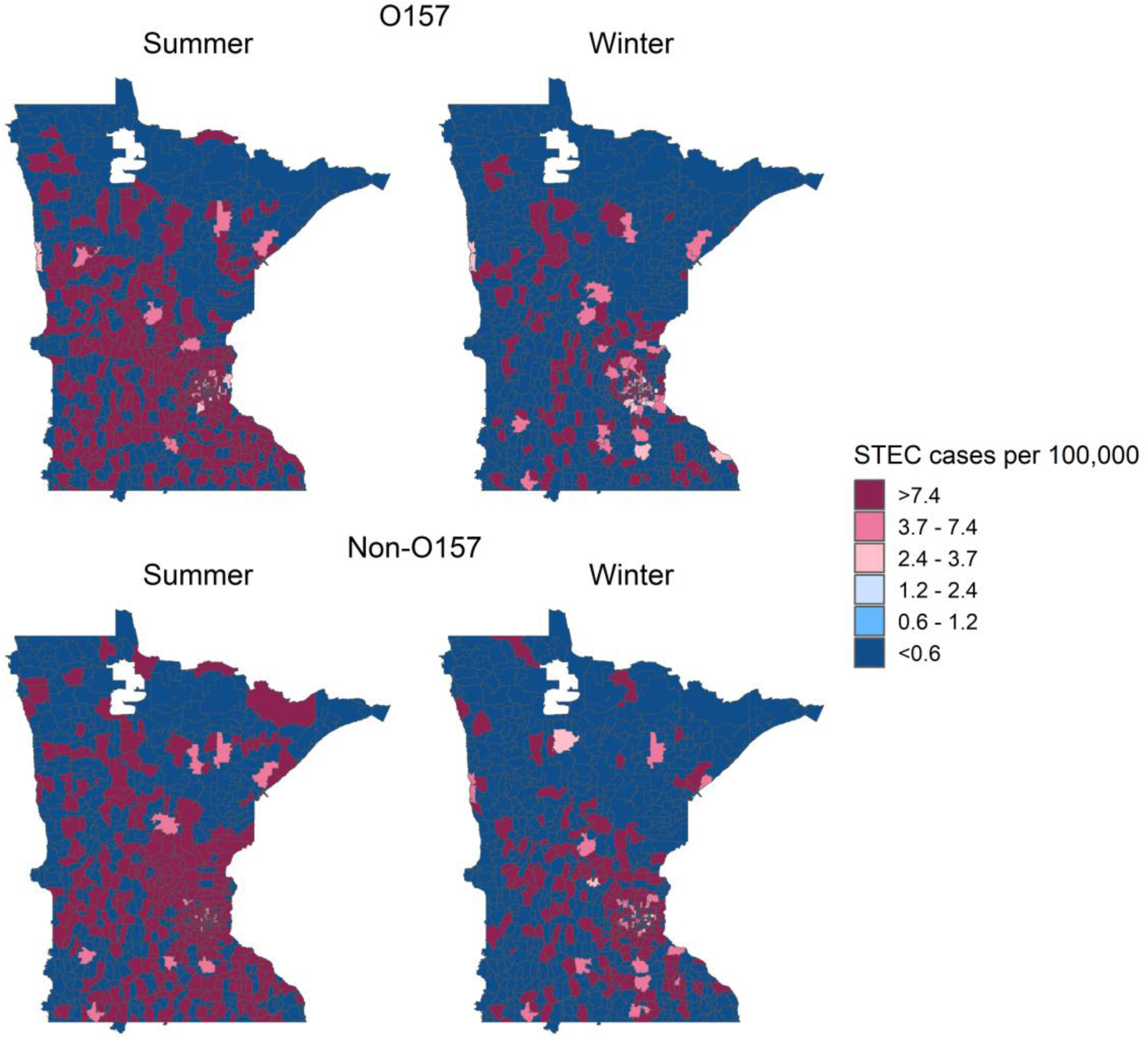
Incidence rate of STEC infections in Minnesota, 2010-2019, by serogroup and season. Incidence is shown as cases per 100,000 population at the zip code tabulation area (ZCTA) level.

Animal density per 10 acres varied spatially across the state (Figure 2). Swine were the most numerous animal we examined and cattle the most common ruminant (Appendix Table A1).

**Figure 2.**
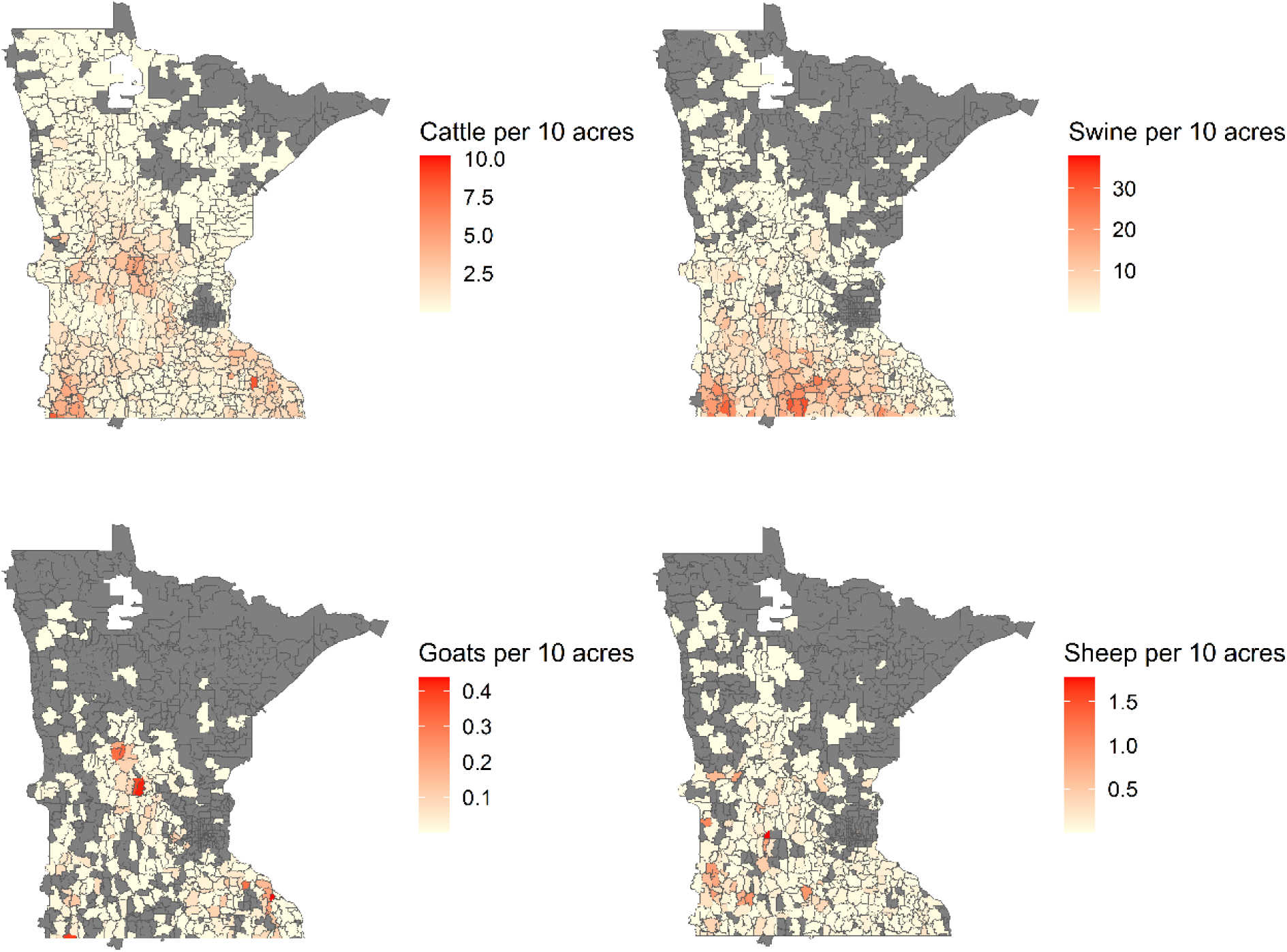
Density in animal operations across Minnesota, covering the period 2010-2019, by animal type. Animal density is shown per 10 acres at the zip code tabulation area (ZCTA) level. ZCTAs without animal operations for a given animal type are shown in grey.

The FoodNet Population Survey collected responses from 3,793 children and adults in Minnesota. We were unable to determine the county of residence for 91 respondents (2.4%), of which 81 had no ZCTA or county reported, and 10 had a ZCTA from outside of Minnesota reported with no county. There were 24 respondents (0.6%) with no data on contact with cows, goats, or sheep in the past 7 days. This left a sample of 3,678 for analysis from 534 of the 881 ZCTAs in Minnesota. Differences in contacts were observed by season, age, and sex (Appendix Table A2). The probability of ruminant contact varied spatially and was greatest among 0-4-year-olds during summer (Figure 3).

**Figure 3.**
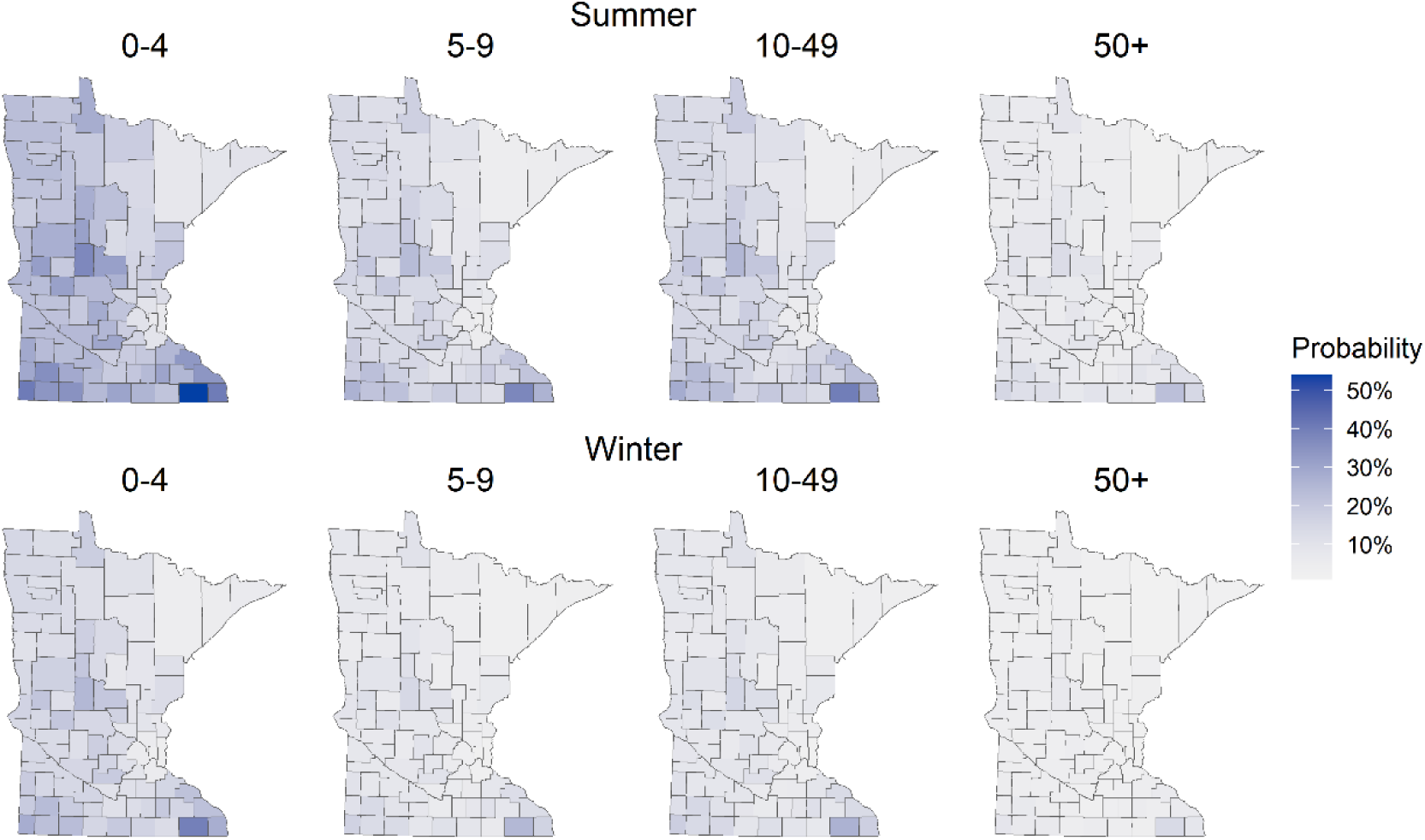
Estimated probability of contact with ruminants in Minnesota, 2010-2019. Probability was calculated from FoodNet Population Survey results from Minnesota with complete data for county and/or zip code and ruminant contact (*29*). Ruminant contact was defined as contact with a cow, sheep, or goat in the previous 7 days.

### Age and Sex Were Associated with Both O157 and Non-O157 STEC

Both age and sex were consistently associated with STEC incidence in Minnesota independent of ruminant density. Regardless of serogroup, season, or adjustment for direct animal contact, 0-4-year-olds were at the greatest risk of STEC infections (Table 2). During summer, the risk decreased with each increasing age group. After adjusting for direct ruminant contact, 5-9-year-olds had a 48% lower rate of O157 STEC (IRR 0.52; 95% CrI 0.41, 0.64) and 59% lower rate of non-O157 STEC (IRR 0.41; 95% CrI 0.31, 0.52) during the summer than 0-4-year-olds. Reductions in incidence were even more pronounced for 10-49 and ≥50-year-olds during the summer (Table 2). During winter, incidence rates were also reduced for individuals ≥5-year-olds across both serogroups, though without the same dose-response relationship as in summer.

**Table 2.** Posterior means and 95% credible intervals for incidence rate ratios of STEC infection by season.

| **Summer** |  |  |  |  |
| --- | --- | --- | --- | --- |
|  | **Without ruminant contact** |  | **With ruminant contact** |  |
| **Variable** | **O157** | **Non-O157** | **O157** | **Non-O157** |
| **Age** |  |  |  |  |
| 0-4 | 1.00 (Ref) | 1.00 (Ref) | 1.00 (Ref) | 1.00 (Ref) |
| 5-9 | 0.44 (0.35, 0.55)<sup>a</sup> | 0.33 (0.26, 0.43)<sup>a</sup> | 0.52 (0.41, 0.64)<sup>a</sup> | 0.41 (0.31, 0.52)<sup>a</sup> |
| 10-49 | 0.24 (0.20, 0.27)<sup>a</sup> | 0.33 (0.28, 0.38)<sup>a</sup> | 0.23 (0.20, 0.27)<sup>a</sup> | 0.32 (0.28, 0.37)<sup>a</sup> |
| 50+ | 0.19 (0.16, 0.23)<sup>a</sup> | 0.15 (0.13, 0.19)<sup>a</sup> | 0.16 (0.13, 0.19)<sup>a</sup> | 0.12 (0.10, 0.15)<sup>a</sup> |
| **Sex** |  |  |  |  |
| Male | 1.00 (Ref) | 1.00 (Ref) | 1.00 (Ref) | 1.00 (Ref) |
| Female | 1.18 (1.04, 1.33)<sup>a</sup> | 1.54 (1.36, 1.74)<sup>a</sup> | 1.14 (1.01, 1.29)<sup>a</sup> | 1.48 (1.31, 1.67)<sup>a</sup> |
| **Cattle per 10 acres** | 1.31 (1.19, 1.44)<sup>a</sup> | 1.02 (0.90, 1.14) | 1.30 (1.18, 1.42)<sup>a</sup> | 0.98 (0.87, 1.10) |
| **Goats per 10 acres** | 3.23 (0.26, 13.69) | 21.52 (1.91, 89.42)<sup>a</sup> | 2.88 (0.23, 12.22) | 19.61 (1.69, 78.84)<sup>a</sup> |
| **Sheep per 10 acres** | 2.35 (1.14, 4.17)<sup>a</sup> | 2.14 (0.95, 3.97) | 2.35 (1.14, 4.20)<sup>a</sup> | 2.15 (0.96, 3.96) |
| **Ruminant contact** |  |  |  |  |
| No | - | - | 1.00 (Ref) | 1.00 (Ref) |
| Yes | - | - | 5.15 (4.51, 5.86)<sup>a</sup> | 7.46 (6.57, 8.43)<sup>a</sup> |
| **Winter** |  |  |  |  |
|  | **Without ruminant contact** |  | **With ruminant contact** |  |
| **Variable** | **O157** | **Non-O157** | **O157** | **Non-O157** |
| **Age** |  |  |  |  |
| 0-4 | 1.00 (Ref) | 1.00 (Ref) | 1.00 (Ref) | 1.00 (Ref) |
| 5-9 | 0.50 (0.29, 0.80)<sup>a</sup> | 0.23 (0.13, 0.37)<sup>a</sup> | 0.55 (0.31, 0.88)<sup>a</sup> | 0.27 (0.15, 0.44)<sup>a</sup> |
| 10-49 | 0.50 (0.36, 0.69)<sup>a</sup> | 0.53 (0.41, 0.68)<sup>a</sup> | 0.49 (0.35, 0.68)<sup>a</sup> | 0.52 (0.41, 0.67)<sup>a</sup> |
| 50+ | 0.46 (0.31, 0.64)<sup>a</sup> | 0.28 (0.21, 0.37)<sup>a</sup> | 0.40 (0.27, 0.56)<sup>a</sup> | 0.22 (0.16, 0.30)<sup>a</sup> |
| **Sex** |  |  |  |  |
| Male | 1.00 (Ref) | 1.00 (Ref) | 1.00 (Ref) | 1.00 (Ref) |
| Female | 1.67 (1.34, 2.07)<sup>a</sup> | 1.64 (1.38, 1.96)<sup>a</sup> | 1.64 (1.31, 2.03)<sup>a</sup> | 1.59 (1.33, 1.89)<sup>a</sup> |
| **Cattle per 10 acres** | 0.96 (0.77, 1.14) | 1.03 (0.87, 1.20) | 0.89 (0.73, 1.07) | 0.98 (0.82, 1.15) |
| **Goats per 10 acres** | 21.04 (0.05, 152.55) | 4.37 (0.03, 28.37) | 21.62 (0.04, 154.68) | 3.98 (0.02, 26.09) |
| **Sheep per 10 acres** | 4.37 (1.49, 9.18)<sup>a</sup> | 2.59 (0.84, 5.51) | 4.28 (1.40, 8.92)<sup>a</sup> | 2.58 (0.82, 5.58) |
| **Ruminant contact** |  |  |  |  |
| No | - | - | 1.00 (Ref) | 1.00 (Ref) |
| Yes | - | - | 4.84 (3.72, 6.17)<sup>a</sup> | 8.86 (7.29, 10.67)<sup>a</sup> |

<sup>a</sup> $p < 0.05$

Individuals reporting female sex were at consistently greater risk of STEC infection, with only slight variation between serogroups and seasons (Table 2). For O157 STEC, the IRR comparing female cases to male ranged from 1.18 (95% CrI 1.04, 1.33) during summer to 1.67 (95% CrI 1.34, 2.07) during winter. Compared to O157 STEC, the relative increase in incidence of non-O157 STEC among female cases was greater during the summer (IRR 1.54; 95% CrI 1.36, 1.73), but did not increase as substantially during winter (IRR 1.64; 95% CrI 1.37, 1.96).

### Direct Ruminant Contact Contributes Substantially to STEC Risk

Direct contact with cattle, sheep, or goats in the 7 days before illness significantly increased the risk of STEC infection across both serogroups and seasons. Among individuals reporting ruminant contact, the observed incidence of O157 STEC infections per 100,000 was 70.9 in summer and 24.5 in winter. For non-O157 STEC infections, incidence per 100,000 was 85.0 in summer and 53.5 in winter. Comparatively, among cases without ruminant contact, the observed O157 STEC incidence rates per 100,000 were 13.6 in summer and 5.3 in winter and for non-O157 STEC incidence were 13.2 in summer and 6.9 in winter.

After adjustment for age, sex, and ruminant density, the effect of direct ruminant contact on O157 STEC incidence was IRR 5.11 (95% CrI 4.47, 5.81) during the summer, and similar during the winter (Table 2). Non-O157 risk increased 7.47-fold (IRR 7.47; 95% CrI 6.58, 8.42) with direct ruminant contact during the summer and 9.01-fold (IRR 9.01; 95% CrI 7.44, 10.83) with direct ruminant contact during the winter. In sensitivity analysis, combining individuals with direct ruminant contact and individuals with only indirect ruminant contact accentuated the effects observed with only direct contact across all serogroups and seasons (Appendix Table A3) For example, during summer, direct and indirect ruminant contact were associated with 7.54 times (IRR 7.54; 95% CrI 6.61, 8.56) the risk of O157 STEC infection and 9.61 times (IRR 9.61; 95% CrI 8.47, 10.9) the risk of non-O157 STEC infection.

### Ruminant Density was Associated with STEC Incidence Independently of Direct Contact

The risk of O157 STEC infection increased for individuals living in a ZCTA with a high density of cattle during summer, and a high density of sheep year-round. (Table 2). During summer, incidence increased 30% for every additional 10 head of cattle (IRR 1.30; 95% CrI 1.18, 1.42) and 135% for every additional 10 sheep (IRR 2.35; 95% CrI 1.14, 4.20). The effect of sheep was greater during winter, with an IRR of 4.28 (95% CrI 1.40, 8.92). Concordantly, O157 STEC rates were greatest in the center and southwest of the state, where large concentrations of cattle and sheep are found (Figure 4).

**Figure 4.**
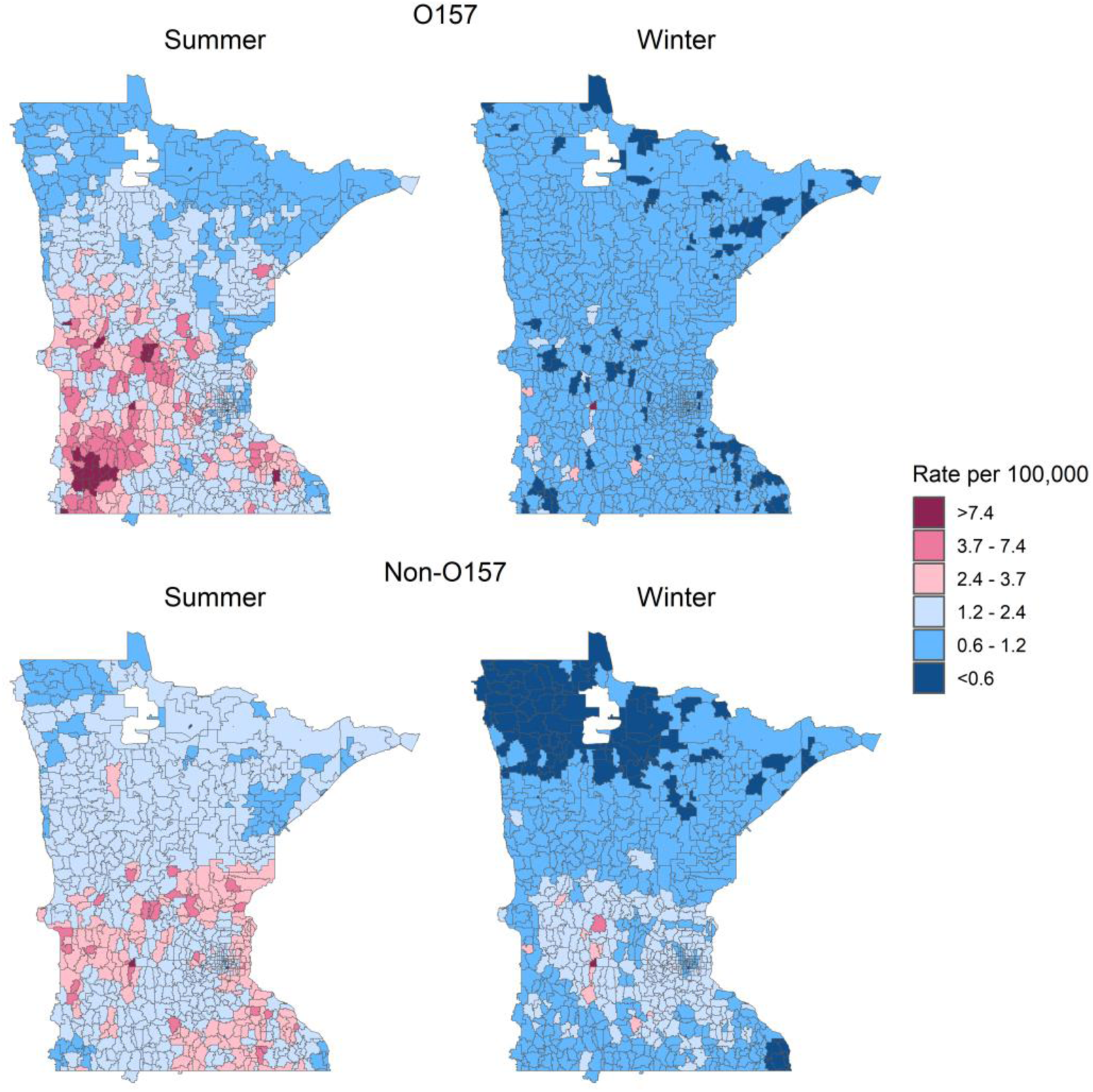
Smoothed incidence rate estimates of Shiga toxin-producing *E. coli* infection by season and serogroup in Minnesota, 2010-2019. Incidence rates were adjusted for age, sex, and direct ruminant contact.

The effects of cattle and sheep density on O157 STEC risk were unaffected by adjustment for direct animal contact (Table 2). The elevated risk during summer persisted in ZCTAs in the central and southwest regions even in the absence of direct animal contact (Figure 5). Conversely, O157 STEC risk during winter was notably low in almost all ZCTAs among individuals without direct ruminant contact, the ZCTA with the greatest density of sheep being the single exception.

**Figure 5.**
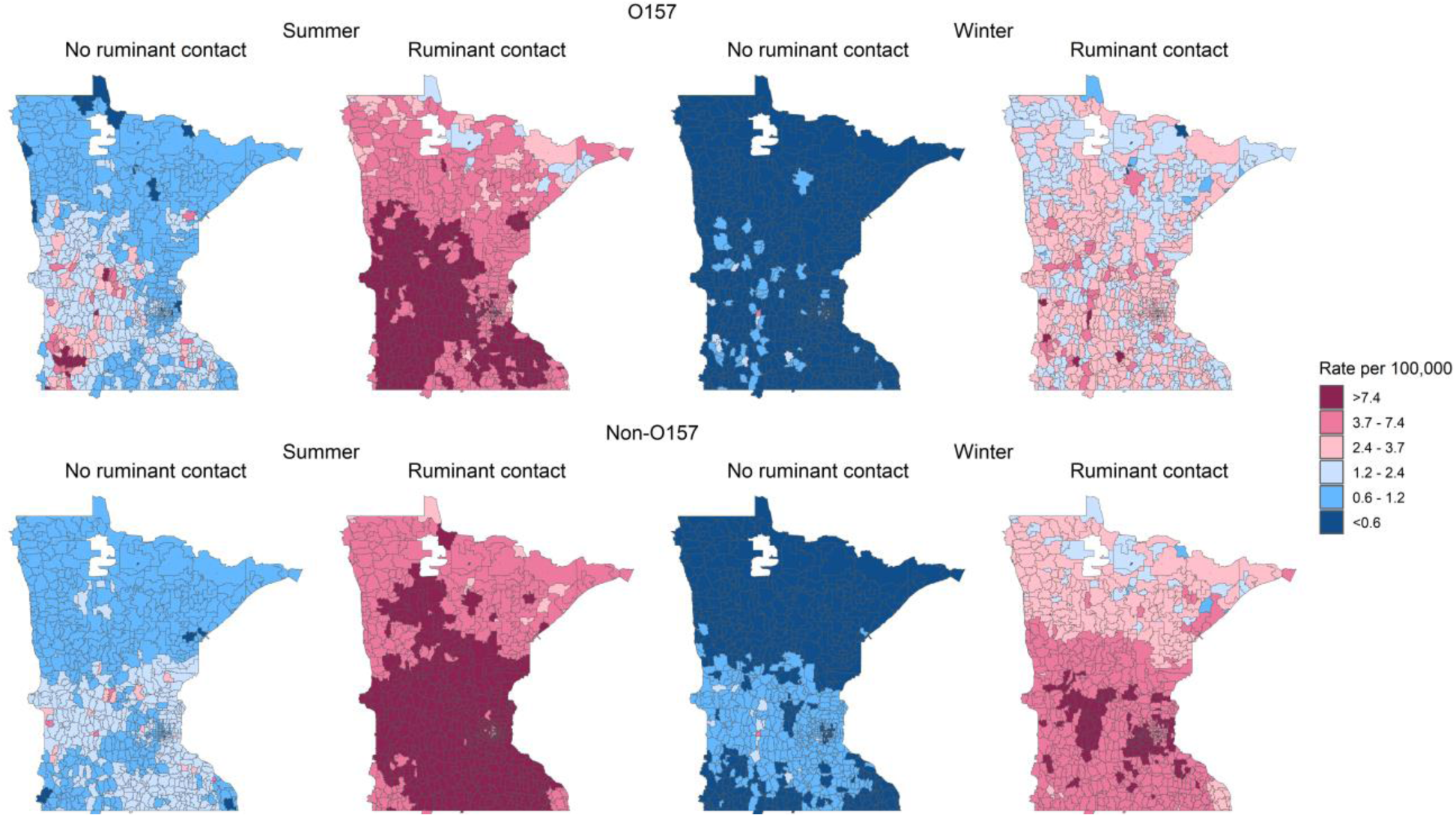
Smoothed incidence rate estimates of Shiga toxin-producing *E. coli* by season, serogroup, and direct ruminant contact in Minnesota, 2010-2019. Incidence rates were adjusted for age and sex.

Non-O157 STEC incidence was associated only with goat density and only during the summer, with an IRR of 19.6 (95% CrI 1.69, 78.8) for each additional goat per 10 acres (Table 2). Ruminant density was not associated with non-O157 STEC incidence during the winter. Non-O157 STEC incidence was greatest in the center and southeast of the state (Figure 4), similar to goat density, though not perfectly aligned (Figure 2). As for O157, the effects of ruminant density on non-O157 STEC incidence did not meaningfully change after adjustment for direct animal contact (Table 2). However, few ZCTAs remained at elevated risk of non-O157 infections in the absence of direct animal contact, though notably the ZCTAs with the highest goat density were among these (Figure 5). Compared to O157 risk, non-O157 incidence was elevated to a greater degree in a greater proportion of ZCTAs during winter.

In our sensitivity analyses, combining individuals with direct ruminant contact with those with only indirect contact made no meaningful different in risk associated with ruminant density (Appendix Table A3), and swine density was not associated with either serogroup in either season (Appendix Table A4).

## Discussion

No prior study has examined the effect of ruminant density on STEC incidence while accounting for direct ruminant contact. We found that the association between living in an area with a high density of ruminants and incident STEC infection was independent of direct ruminant contact for both O157 and non-O157 STEC, in both summer and winter. Our study also demonstrates important differences in the associations of O157 and non-O157 STEC with agricultural animal reservoirs. Cattle and sheep density were specifically associated with O157 STEC incidence in summer and year-round, respectively, and goat density was associated with non-O157 incidence in summer. The risk associated with ruminant density was largely unaffected by the inclusion of direct or indirect ruminant contact, which significantly increased risk of both serogroups in both seasons.

High ruminant density can theoretically increase the risk of STEC through multiple mechanisms. Direct animal contact is principal among these, as shown by our stratified maps of ZCTA incidence. During summer, for the population with direct ruminant contact, we found that incidence rates of O157 and non-O157 STEC infections in almost all ZCTAs were each >3.7/100,000, the Healthy People 2030 target for all serogroups combined (35). Even during winter, at least half the ZCTAs had incidence rates >2.4/100,000 among the population with direct ruminant contact. This is consistent with the estimated 11% of STEC outbreaks caused by direct animal contact in the U.S. from 2010-2017 (19) and previous studies showing that direct contact with cattle, particularly, has been associated with increased STEC risk (20–22).

We also assessed the role of indirect contact through exposure to ruminant environments. When we examined the effect of any ruminant contact, direct or indirect, the risk of O157 and non-O157 STEC infection was greater (e.g., IRR 7.5 for the summer, O157 analysis) than when incorporating only direct contact (e.g., IRR 5.1). This suggests substantial STEC transmission occurring when an individual lives in, works in, or visits areas with ruminants, even without coming into direct contact with the animals. Contamination in soil around animal pens and on fences or other fomites can pose an ongoing risk of STEC exposure, as demonstrated by an O157 outbreak among children visiting a barn over a week after it had held cattle (37). Secondary transmission can also occur from individuals infected with STEC acquired through direct ruminant contact. Vigilance in hand hygiene among individuals exposed to ruminant environments but without direct contact and thorough decontamination of spaces previously inhabited by ruminants could help reduce the risk of STEC infections associated with indirect transmission.

Direct or indirect contact is not the only way in which living in a ruminant-dense area can increase STEC risk. Our analysis demonstrates the important role of residual environmental risk from ruminants in a region, independent of direct or indirect ruminant contact. The insensitivity of ruminant density risk to adjustment for ruminant contact is likely due to the only partial overlap of ruminant-dense ZCTAs (Figure 2) and ZCTAs with substantial direct ruminant contact (Figure 3). The risk of direct ruminant contact was driven by statewide patterns and not disproportionately by ZCTAs with high ruminant densities, so it had minimal impact on the risks associated with ruminant density. We found that ZCTAs with STEC infection risk >2.4/100,000 in the absence of direct contact were almost exclusively those with a high density of ruminants (Figure 5).

The residual environmental risk implied by the associations with ruminant density is likely composed of multiple transmission routes, including via locally produced ruminant food products, person-to-person (e.g., in child care or household settings) from an individual who had direct contact with a ruminant, and via environmental reservoirs such as water contaminated by ruminants. Even dust contaminated by STEC has been shown to drift out of ruminant environments to contaminate neighboring areas (38). Transmission chains involving other animal species as intermediaries and direct contact with them could also be involved. How each of these pathways contributes to the risk of living in a region with high ruminant density is unknown and an area of ongoing work. Although no previous study has attempted to disentangle ruminant contact from residual environmental risk, in Scotland, 26% of cases were estimated to be environmental in origin (6), which would include both direct and indirect animal contact and residual environmental risk.

Cattle are the prototypic STEC reservoir, largely because of their role in maintaining and transmitting O157 STEC. Our finding that O157 is associated with cattle density is consistent with several previous studies, including assessments of both ecological (8–13), and direct contact (20–22) risk. Our estimate of the magnitude of the association, IRR 1.30 for each additional head of cattle per 10 acres, was almost identical to that estimated by Frank et al. in Germany for O157 (2.45 per 100 cattle/km^2^, or ∼1.36 per 1 cattle/10 acres) (11). Friesema et al. also stratified by season and similarly found an association between cattle density and O157 STEC incidence only during the summer months (12). However, Mulder et al. observed an association of O157 STEC infections and small ruminant but not cattle density, and when small ruminants were analyzed separately, only goats, not sheep, were associated with O157 incidence (14). This inconsistency could be due to differences in local STEC ecology between Minnesota and the Netherlands. With a different array of STEC strains, hosts may also differ. Our findings demonstrate significant residual environmental O157 risk associated with cattle density and indicate transmission pathways from cattle to the public, including through food products and environmental contamination, should to be identified at a local level for tailored public health prevention measures.

We found that infection with the O157 serogroup was also associated with sheep density, which agrees with other studies that have found an association between O157 and both cattle and sheep (6, 10). Moreover, genomic evidence suggests that O157 strains circulate interchangeably between cattle and sheep (18), supporting the finding that both cattle and sheep density would contribute to STEC incidence. Sheep also stood out as the only species to be associated with STEC incidence during what we defined as the “winter” months, November-April. Among individuals without ruminant contact during winter, the only ZCTA with >2.4/100,000 O157 and non-O157 STEC cases was the ZCTA with the highest sheep density. This may be because sheering and lambing often occur in March and April, involving greater person-sheep contact than at other times of the year.

Surprisingly, we found that non-O157 incidence was not associated with cattle density. Studies from both Europe and North America have identified high levels of non-O157 STEC carriage among cattle (39–42), and living, working, or visiting a farm or public animal venue has been associated with increased risk of non-O157 infection (43). However, only one other study, conducted in Germany, has examined the risk of cattle density on non-O157 STEC incidence. In that study, they found that cattle density was associated with increased incidence of O111, O103, and O145, but not O26 (11). It is possible that if we had sufficient power to examine individual non-O157 serogroups, we would have identified this type of heterogeneity; however, it is also possible that adjusting for the presence of other animals, particularly sheep and goats, would have nullified the associations they observed for cattle, as agricultural areas are likely to contain multiple species. Geographic variation in reservoirs is also likely, and cattle may not be as significant a source of non-O157 STEC infections in Minnesota as they were in Germany.

Non-O157 STEC have been associated with visiting petting zoos (43, 44), where goats and sheep are the two most common species available (45). At 19.5 after adjustment for direct animal contact, the IRR we estimated for the association of goats and non-O157 STEC incidence was the highest we observed. The ZCTA with the highest density of goats had 0.44 goats per 10 acres, and the IRR quantifies the increase in risk for the addition of 1 goat per 10 acres. Thus, even in the highest density ZCTA, the addition of 1 goat/10 acres would be equivalent to more than tripling the number of goats. If goats pose a risk of non-O157 STEC infection, such a relatively large increase can be understood to have the outsized impact on risk that we observed. However, the high IRR we estimated is also not unprecedented. In one case-control study, contact with goats was associated with a 21-fold increase in the odds of non-O157 STEC infection (43). Goats may be an important reservoir of non-O157 STEC in Minnesota, and their popularity in agritourism suggests that additional prevention measures may be warranted at public animal contact venues.

Although STEC are most commonly found among ruminant reservoirs, outbreaks in Canada have been linked to pork products (46, 47), and at least one study has found an increased risk of non-O157 STEC infection associated with living on a farm with swine (43). After accounting for the presence of ruminant reservoirs, however, our study did not detect an association between STEC incidence and swine density. While swine are competent STEC hosts, they do not appear to serve as an important source of residual environmental risk for STEC infections in Minnesota.

While similar patterns of STEC risk by age and sex have been observed before, the literature is inconsistent in the age groups used for analysis, and an effect of sex has only been observed in some studies (12, 48–52). We found that 0-4 year olds were at greatest risk of both O157 and non-O157 STEC, independent of ruminant density and contact, which is consistent with previous studies showing 0-4 or 1-4 year olds at greatest risk of infection (12, 49, 53). Elevated risk in young children may be from greater exposure to STEC, reporting bias due to increased severity and lower thresholds for healthcare-seeking and diagnostic testing seen in this age group, or naïve immune systems. The dose-response relationship between greater age and lower STEC risk during the summer is suggestive of higher thresholds for healthcare-seeking and diagnostic testing as age increases or the acquisition of immunity over time, which has been reported previously in studies of farmworkers and their families (54, 55). We also found that during summer, independent of ruminant density and contact, the relative increase in non-O157 STEC risk for cases reporting female sex was greater than the increase in O157 STEC, and the highest IRRs across serogroups were observed during winter. This likely indicates a relative increase in transmission routes more common among individuals of female sex during the winter, and that such transmission routes are more important for non-O157 than O157 STEC.

The ZCTAs with the greatest STEC risk were overwhelmingly rural, consistent with previous work (6, 10, 13). Understanding the source of STEC infections in rural areas, including direct and indirect animal contact, secondary transmission, well water, and environmental contamination, is central to prevention efforts. Health care access can be more difficult in rural areas, particularly with hospital closures (56, 57), which may put STEC cases in rural areas at greater risk of severe outcomes such as HUS. With O157 STEC still the largest cause of HUS, prevention in rural areas is a priority.

Our study was limited by insufficient sample size to analyze individual non-O157 serogroups. While there is likely some heterogeneity in reservoirs between serogroups, a previous study found the effect of ruminant density to be mostly consistent across non-O157 serogroups (11). The Minnesota Pollution Control Agency’s definition of a feedlot excluded operations with <50 animals (or <10 animals within shorelands), so our analysis does not include small farms. We believe the impact of this is minimal, as small farms are likely to be located in the same regions as larger operations. To incorporate individual-level direct ruminant contact, we had to determine what portion of the population is exposed to ruminants. The FoodNet Population Survey we used to do this asked about contact with cows, sheep, or goats in a single question, prohibiting us from assessing the effect of direct contact with individual species. The FoodNet Population Survey was also limited in its sample size, leaving the populations in 347 of 881 ZCTAs unsurveyed, which required us to determine the expected number of individuals with ruminant contact at the county level. Consequently, ZCTA-level expected contact counts were not as specific as they would have been if all ZCTAs had been covered by the survey. Finally, unrecognized factors may have confounded these results. To minimize this possibility, we adjusted for age and sex, jointly modeled all species in our analysis, and accounted for two types of spatial correlation.

### Conclusion

Our results indicate a need to identify and mitigate transmission routes from local cattle, sheep, and goat reservoir populations. For the first time, our study demonstrates that the risk posed by living in an area with high ruminant density does not operate solely through direct contact with ruminants or even exposure to ruminant environments. Thus, more work is needed to identify prevention measures for local transmission occurring through food, person-to-person contact with individuals who encounter ruminants, and contamination of neighboring areas including water bodies and produce fields. At the same time, both direct and indirect ruminant contact dramatically increase an individual’s risk of O157 and non-O157 STEC at all times of year, emphasizing the importance of reinforcing handwashing and other best practices for contacting ruminants.

## Acknowledgements

This research was funded in part by NIH/NIAID K01AI168499. The funder had no role in study design, data collection and interpretation, or the decision to submit the work for publication. We gratefully acknowledge the use of data from the 2018-2019 FoodNet Population Survey, supplied by CDC.

## Author Contributions

CW: Data curation, Formal analysis, Methodology, Software, Validation, Visualization, Writing – original draft; WF: Data curation, Writing – original draft; KS: Conceptualization, Investigation, Resources, Writing – review & editing; JR: Investigation, Resources, Writing – review & editing; CAK: Investigation, Writing – review & editing; GAMT: Conceptualization, Funding acquisition, Methodology, Writing – original draft.

## Data Availability

Data on individuals with reported STEC infections is available upon request and with an appropriate agreement from the Minnesota Department of Health. Analysis code is available upon request from the corresponding author.

## Appendix Tables

**Table A1.** Characteristics of reported STEC cases, Minnesota 2010-2019, by serogroup. Unknown values were imputed prior to analysis.

| **Animal Type** | **Number of Feedlots** | **Number of Animals** | **Mean per Feedlot <sup>a</sup>** |
| --- | --- | --- | --- |
| **Cattle** | 18,441 | 2,967,940 | 160.9 |
| **Swine** | 5,869 | 10,611,608 | 1808.1 |
| **Goats** | 684 | 45,262 | 66.2 |
| **Sheep** | 1,347 | 206,509 | 153.3 |

<sup>a</sup> Mean animals per feedlot calculated among feedlots with >0 animals of the given type.

**Table A2.** Estimated percentages of individuals with contact with a cow, sheep, or goat in the past 7 days by season, age, and sex, calculated from the FoodNet Population Survey (*29*).

| Season | Age | Male (%) | Female (%) | Both Sexes Combined (%) |
| --- | --- | --- | --- | --- |
| **Summer** | **0-4** | 14.40 (14.01, 14.81) | 16.05 (8.58, 28.03) | 15.29 (10.83, 21.14) |
|  | **4-9** | 7.49 (5.51, 10.10) | 11.12 (5.74, 20.45) | 9.05 (7.03, 11.57) |
|  | **10-49** | 3.68 (2.46, 5.46) | 12.14 (9.9, 14.81) | 8.22 (6.44, 10.45) |
|  | **50+** | 3.74 (3.22, 4.36) | 5.59 (3.87, 8.01) | 4.69 (3.98, 5.52) |
| **Winter** | **0-4** | 3.62 (2.23, 5.83) | 6.79 (4.59, 9.93) | 5.03 (3.23, 7.75) |
|  | **4-9** | 3.07 (2.33, 4.06) | 10.10 (6.88, 14.6) | 6.88 (4.92, 9.55) |
|  | **10-49** | 4.35 (3.51, 5.39) | 8.68 (6.64, 11.28) | 6.38 (5.72, 7.12) |
|  | **50+** | 4.73 (3.94, 5.67) | 3.54 (2.73, 4.58) | 4.11 (3.54, 4.77) |

**Table A3.** Posterior means and 95% credible intervals for incidence rate ratios of STEC infection, combining direct ruminant contact and any reported exposure to a ruminant environment.

| **Summer** |  |  |  |  |
| --- | --- | --- | --- | --- |
|  | **With direct ruminant contact only** |  | **With direct + indirect ruminant contact** |  |
| **Variable** | **O157** | **Non-O157** | **O157** | **Non-O157** |
| **Age** |  |  |  |  |
| 0-4 | 1.00 (Ref) | 1.00 (Ref) | 1.00 (Ref) | 1.00 (Ref) |
| 5-9 | 0.52 (0.41, 0.64)<sup>a</sup> | 0.41 (0.31, 0.52)<sup>a</sup> | 0.55 (0.43, 0.68)<sup>a</sup> | 0.43 (0.33, 0.55)<sup>a</sup> |
| 10-49 | 0.23(0.20, 0.27)<sup>a</sup> | 0.32 (0.28, 0.37)<sup>a</sup> | 0.23 (0.20, 0.27)<sup>a</sup> | 0.32 (0.28, 0.37)<sup>a</sup> |
| 50+ | 0.16 (0.13, 0.19)<sup>a</sup> | 0.12 (0.10, 0.15)<sup>a</sup> | 0.15 (0.12, 0.18)<sup>a</sup> | 0.12 (0.09, 0.14)<sup>a</sup> |
| **Sex** |  |  |  |  |
| Male | 1.00 (Ref) | 1.00 (Ref) | 1.00 (Ref) | 1.00 (Ref) |
| Female | 1.18 (1.04, 1.33)<sup>a</sup> | 1.54 (1.36, 1.73)<sup>a</sup> | 1.18 (1.04, 1.33)<sup>a</sup> | 1.54 (1.36, 1.73)<sup>a</sup> |
| **Cattle per 10 acres** | 1.30 (1.18, 1.42)<sup>a</sup> | 0.98 (0.87, 1.11) | 1.29 (1.17, 1.41)<sup>a</sup> | 0.98 (0.86, 1.10) |
| **Goats per 10 acres** | 2.84 (0.23, 11.96) | 19.51 (1.66, 79.08)<sup>a</sup> | 2.78 (0.22, 11.70) | 19.32 (1.70, 78.21)<sup>a</sup> |
| **Sheep per 10 acres** | 2.36 (1.14, 4.17)<sup>a</sup> | 2.15 (0.95, 4.00)<sup>a</sup> | 2.35 (1.12, 4.15)<sup>a</sup> | 2.14 (0.94, 3.99) |
| **Ruminant contact** |  |  |  |  |
| No | 1.00 (Ref) | 1.00 (Ref) | 1.00 (Ref) | 1.00 (Ref) |
| Yes | 5.11 (4.47, 5.81)<sup>a</sup> | 7.47 (6.58, 8.42)<sup>a</sup> | 7.54 (6.61, 8.56)<sup>a</sup> | 9.61 (8.47, 10.88)<sup>a</sup> |
| **Winter** |  |  |  |  |
|  | **With direct ruminant contact only** |  | **With direct + indirect ruminant contact** |  |
| **Variable** | **O157** | **Non-O157** | **O157** | **Non-O157** |
| **Age** |  |  |  |  |
| 0-4 | 1.00 (Ref) | 1.00 (Ref) | 1.00 (Ref) | 1.00 (Ref) |
| 5-9 | 0.55 (0.31, 0.87)<sup>a</sup> | 0.27 (0.16, 0.43)<sup>a</sup> | 0.56 (0.32, 0.90)<sup>a</sup> | 0.28 (0.16, 0.44)<sup>a</sup> |
| 10-49 | 0.50 (0.35, 0.68)<sup>a</sup> | 0.52 (0.41, 0.66)<sup>a</sup> | 0.50 (0.35, 0.68)<sup>a</sup> | 0.52 (0.41, 0.66)<sup>a</sup> |
| 50+ | 0.40 (0.27, 0.56)<sup>a</sup> | 0.22 (0.16, 0.29)<sup>a</sup> | 0.38 (0.27, 0.54)<sup>a</sup> | 0.22 (0.16, 0.29)<sup>a</sup> |
| **Sex** |  |  |  |  |
| Male | 1.00 (Ref) | 1.00 (Ref) | 1.00 (Ref) | 1.00 (Ref) |
| Female | 1.67 (1.34, 2.07)<sup>a</sup> | 1.64 (1.37, 1.96)<sup>a</sup> | 1.68 (1.34, 2.08)<sup>a</sup> | 1.64 (1.38, 1.95)<sup>a</sup> |
| **Cattle per 10 acres** | 0.89 (0.73, 1.07) | 0.98 (0.82, 1.15) | 0.87 (0.71, 1.05) | 0.98 (0.82, 1.15) |
| **Goats per 10 acres** | 20.03 (0.04, 137.85) | 3.79 (0.02, 24.90) | 20.03 (0.04, 137.85)<sup>a</sup> | 3.86 (0.02, 24.62) |
| **Sheep per 10 acres** | 4.29 (1.42, 8.92)<sup>a</sup> | 2.59 (0.84, 5.66) | 4.29 (1.42, 8.92)<sup>a</sup> | 2.59 (0.83, 5.62) |
| **Ruminant contact** |  |  |  |  |
| No | 1.00 (Ref) | 1.00 (Ref) | 1.00 (Ref) | 1.00 (Ref) |
| Yes | 4.93 (3.76, 6.32)<sup>a</sup> | 9.01 (7.44, 10.83)<sup>a</sup> | 6.66 (5.20, 8.40)<sup>a</sup> | 10.70 (8.87, 12.81)<sup>a</sup> |

<sup>a</sup> $p < 0.05$

**Table A4.** Posterior means and 95% credible intervals for incidence rate ratios of STEC infection, including swine density.

|  |  | Without ruminant contacts |  | With ruminant contacts |  |
| --- | --- | --- | --- | --- | --- |
|  | Variable | O157 | Non-O157 | O157 | Non-O157 |
| Summer | Age |  |  |  |  |
|  | 0-4 | 1.00 (ref) | 1.00 (ref) | 1.00 (ref) | 1.00 (ref) |
|  | 5-9 | 0.44 (0.35, 0.55) | 0.33 (0.26, 0.42) | 0.52 (0.41, 0.64) | 0.41 (0.31, 0.52) |
|  | 10-49 | 0.24 (0.20, 0.28) | 0.33 (0.28, 0.38) | 0.23 (0.20, 0.27) | 0.32 (0.28, 0.37) |
|  | 50+ | 0.19 (0.16, 0.23) | 0.15 (0.13, 0.19) | 0.16 (0.13, 0.19) | 0.12 (0.10, 0.15) |
|  | Sex |  |  |  |  |
|  | Male | 1.00 (ref) | 1.00 (ref) | 1.00 (ref) | 1.00 (ref) |
|  | Female | 1.18 (1.04, 1.33) | 1.54 (1.36, 1.73) | 1.18 (1.04, 1.33) | 1.54 (1.36, 1.73) |
|  | Cattle per 10 acres | 1.31 (1.19, 1.44) | 1.02 (0.90, 1.14) | 1.30 (1.18, 1.42) | 0.99 (0.87, 1.11) |
|  | Goats per 10 acres | 3.31 (0.25, 14.32) | 21.22 (1.85, 88.68) | 2.86 (0.22, 12.04) | 19.42 (1.67, 79.69) |
|  | Sheep per 10 acres | 2.36 (1.14, 4.20) | 2.14 (0.94, 3.95) | 2.36 (1.13, 4.20) | 2.16 (0.95, 4.01) |
|  | Swine per 10 acres | 1.00 (0.97, 1.03) | 1.00 (0.97, 1.03) | 1.00 (0.97, 1.03) | 1.00 (0.96, 1.03) |
|  | Ruminant contact |  |  |  |  |
|  | No | 1.00 (ref) | 1.00 (ref) | 1.00 (ref) | 1.00 (ref) |
|  | Yes | - | - | 5.10 (4.46, 5.81) | 7.47 (6.58, 8.45) |
| Winter | Age |  |  |  |  |
|  | 0-4 | 1.00 (ref) | 1.00 (ref) | 1.00 (ref) | 1.00 (ref) |
|  | 5-9 | 0.50 (0.29, 0.81) | 0.23 (0.13, 0.37) | 0.55 (0.31, 0.88) | 0.27 (0.15, 0.43) |
|  | 10-49 | 0.50 (0.36, 0.69) | 0.53 (0.41, 0.68) | 0.50 (0.35, 0.68) | 0.52 (0.41, 0.67) |
|  | 50+ | 0.46 (0.32, 0.65) | 0.28 (0.20, 0.37) | 0.40 (0.28, 0.56) | 0.22 (0.16, 0.30) |
|  | Sex |  |  |  |  |
|  | Male | 1.00 (ref) | 1.00 (ref) | 1.00 (ref) | 1.00 (ref) |
|  | Female | 1.67 (1.34, 2.07) | 1.64 (1.38, 1.95) | 1.68 (1.34, 2.07) | 1.65 (1.38, 1.96) |
|  | Cattle per 10 acres | 0.97 (0.78, 1.16) | 1.04 (0.87, 1.22) | 0.91 (0.74, 1.10) | 1.00 (0.83, 1.17) |
|  | Goats per 10 acres | 19.42 (0.05, 136.53) | 4.18 (0.03, 26.50) | 18.10 (0.04, 128.20) | 3.72 (0.02, 24.22) |
|  | Sheep per 10 acres | 4.49 (1.45, 9.41) | 2.65 (0.86, 5.64) | 4.57 (1.49, 9.62) | 2.66 (0.83, 5.75) |
|  | Swine per 10 acres | 0.98 (0.94, 1.03) | 0.98 (0.94, 1.03) | 0.98 (0.94, 1.02) | 0.98 (0.94, 1.03) |
|  | Ruminant contact |  |  |  |  |
|  | No | 1.00 (ref) | 1.00 (ref) | 1.00 (ref) | 1.00 (ref) |
|  | Yes | - | - | 4.94 (3.77, 6.31) | 9.02 (7.42, 10.82) |

